# Tracing SARS-CoV-2 Clusters Across Local Scales Using Genomic Data

**DOI:** 10.1101/2024.09.18.24313896

**Authors:** Leke Lyu, Mandev Gill, Guppy Stott, Sachin Subedi, Cody Dailey, Gabriella Veytsel, Magdy Alabady, Kayo Fujimoto, Ryker Penn, Pamela Brown, Roger Sealy, Justin Bahl

**Affiliations:** Center for Ecology of Infectious Diseases, Institute of Bioinformatics, Department of Infectious Diseases, Department of Epidemiology and Biostatistics, University of Georgia, Athens, GA, USA; Department of Statistics, Institute of Bioinformatics, Center for Ecology of Infectious Diseases, University of Georgia, Athens, GA, USA; Georgia Genomics and Bioinformatics Center, University of Georgia, Athens, GA, USA; Department of Health Promotion and Behavioral Sciences, The University of Texas Health Science Center at Houston, Houston, TX, USA; Houston Health Department, Houston, TX, USA

**Keywords:** Viral evolution, Genomic epidemiology, Pandemic control

## Abstract

Quantitatively understanding local transmission dynamics is essential for designing effective prevention strategies. In this study, we developed a novel algorithm to identify introductions and trace locally circulating clusters. We analyzed over 26,000 SARS-CoV-2 genomes and their associated metadata, collected between January and October 2021, to explore introduction and dispersal patterns in Greater Houston, a major metropolitan area known for its demographic diversity. Our analysis identified more than 1,000 independent introduction events, resulting in clusters of varying sizes. Earlier clusters were generally larger and posed greater challenges for control efforts. Characterization of introduction sources revealed that domestic origins were more significant than international ones. Additionally, analysis of locally circulating clusters highlighted age-structured transmission dynamics. Geographic reconstruction of cluster spread identified Harris County as the primary viral source for surrounding counties. Harris county sustained the local epidemic with a smaller proportion of new cases driven by external importations and longer persistence times of circulating lineages. Overall, our high-resolution spatiotemporal reconstruction of the epidemic in Greater Houston provides critical insights into the heterogeneous transmission landscape, supporting regional response strategies and public health planning.

**Significance Statement:** The growing recognition of genome sequencing as a critical tool for outbreak response has driven a rapid increase in the availability of sequence data. Here, we present an analytical workflow to trace imported SARS-CoV-2 clusters using large-scale genome datasets. Our approach pinpoints when, where, and how many introductions occurred, while also tracking the circulation of resulting clusters. By incorporating metrics such as the Source Sink Score, Local Import Score, and Persistence Time, our analysis reveals transmission heterogeneity between subregions of the focal area. These insights are essential for monitoring viral introductions and guiding targeted control measures, enhancing the ability of local responders to address the challenges of current and future pandemics as new variants emerge.

## Introduction

Genome epidemiology has significantly advanced our understanding of and efforts to combat emerging infectious diseases (1–3). In the context of SARS-CoV-2, previous research has demonstrated its capability to clarify the virus’s origins and spread (4, 5), reconstruct local transmission chains (6), assess the effectiveness of non-pharmaceutical interventions (7), and identify key predictors of viral lineage movements (8). These epidemiological insights, translated from the virus’s evolutionary history, are crucial for shaping public health policies and would not have been possible without extensive sequencing efforts. By August 2024, over 16 million genome sequences had been submitted to the Global Initiative on Sharing All Influenza Data (GISAID) (9). Although these expansive COVID-19 datasets facilitate high-resolution inferences about local transmission dynamics, they also present significant computational challenges (10). In response, new algorithms, software, and computational workflows have emerged since the onset of the pandemic, including tools for rapid phylogenetic tree construction (11, 12) and the Thorney BEAST module for more efficient generation of time-resolved trees (13, 14).

Houston, the largest city in the Southern United States, anchors the Greater Houston metropolitan area and is one of the most demographically diverse cities in the country (15). It is also one of the most economically segregated cities, marked by sharp divisions in income, education, and occupation (16). According to Covid Act Now, Houston faces considerable challenges due to its high population density, significant proportion of non-English speakers, and notable income disparity. These factors strain the city’s health system, making Houston more vulnerable to a SARS-CoV-2 outbreak than over 90% of U.S. metropolitan areas (17). This vulnerability highlights the urgent need to understand local patterns of SARS-CoV-2 dispersal and how these patterns vary across different socioeconomic regions.

The COVID-19 pandemic has been characterized by the emergence and spread of genetically distinct virus variants that exhibit increased transmissibility compared to earlier lineages (18). The Delta variant, in particular, not only spreads more rapidly (19) but also leads to higher rates of hospitalization (20) and demonstrates greater immune evasion (21) than the previously dominant Alpha variant. In Houston, the emergence of Delta occurred in a context of heterogeneous prior immunity, resulting from both previous infections and vaccinations. The first case of the Delta variant in Houston Methodist Hospital was identified in mid-April 2021, during a period of declining COVID-19 cases (22). Beginning in early July, there was a sharp increase in cases driven by the Delta variant, with these cases doubling in frequency approximately every seven days (22). Several critical questions remain unanswered, which cannot be resolved without genomic epidemiological inference. What is the primary source of these variants? What role did Houston play in the introduction of Delta to the U.S.?

With the support of the Houston Health Department (HHD), we accessed an extensive dataset comprising over 10,000 Delta genomes sampled from Houston between January 2021 and October 2021, each linked with metadata such as zip code, age, and sex. This dataset offers a valuable opportunity to investigate transmission dynamics in Houston, examine how population structure influences disease spread, and assess variations in SARS-CoV-2 transmission across different subregions.

A notable phylodynamic workflow developed by Dellicour et al. (23) facilitates large-scale phylogeographic analysis through two principal steps: first, a preliminary discrete trait analysis (24) on fixed empirical topologies identifies introduction events; second, it estimates the circulation dynamics of local viral clusters (25, 26). We adapted this analytical workflow to examine the spatial invasion dynamics of SARS-CoV-2 in Greater Houston, as illustrated in Figure S1. Applying a novel algorithm, we determined the timing and number of viral introductions and tracked the resulting circulating clusters. We specifically investigated whether international or domestic introductions played a more prominent role. Additionally, we modeled the transmission structure among different demographic groups, determined by sex and age. Finally, we explored the spatiotemporal variation in viral dispersal across various subregions, defined here as counties, within Greater Houston.

## Results

### Identifying Distinct Introduction Events and Locally Circulating Clusters

Our dataset comprised 26,138 complete SARS-CoV-2 genomes, including 9,186 sampled from Houston and 16,952 contextual sequences from around the world. We conducted a discrete phylogeographic reconstruction on the time-calibrated phylogeny. Considering ‘Houston’ and ‘non-Houston’ as the two possible locations, introduction events were identified on branches where a state change occurred, specifically from ‘non-Houston’ to ‘Houston.’ Subtrees extending from these introduction events were classified as locally circulating clusters (Figure 1).

**Figure 1.**
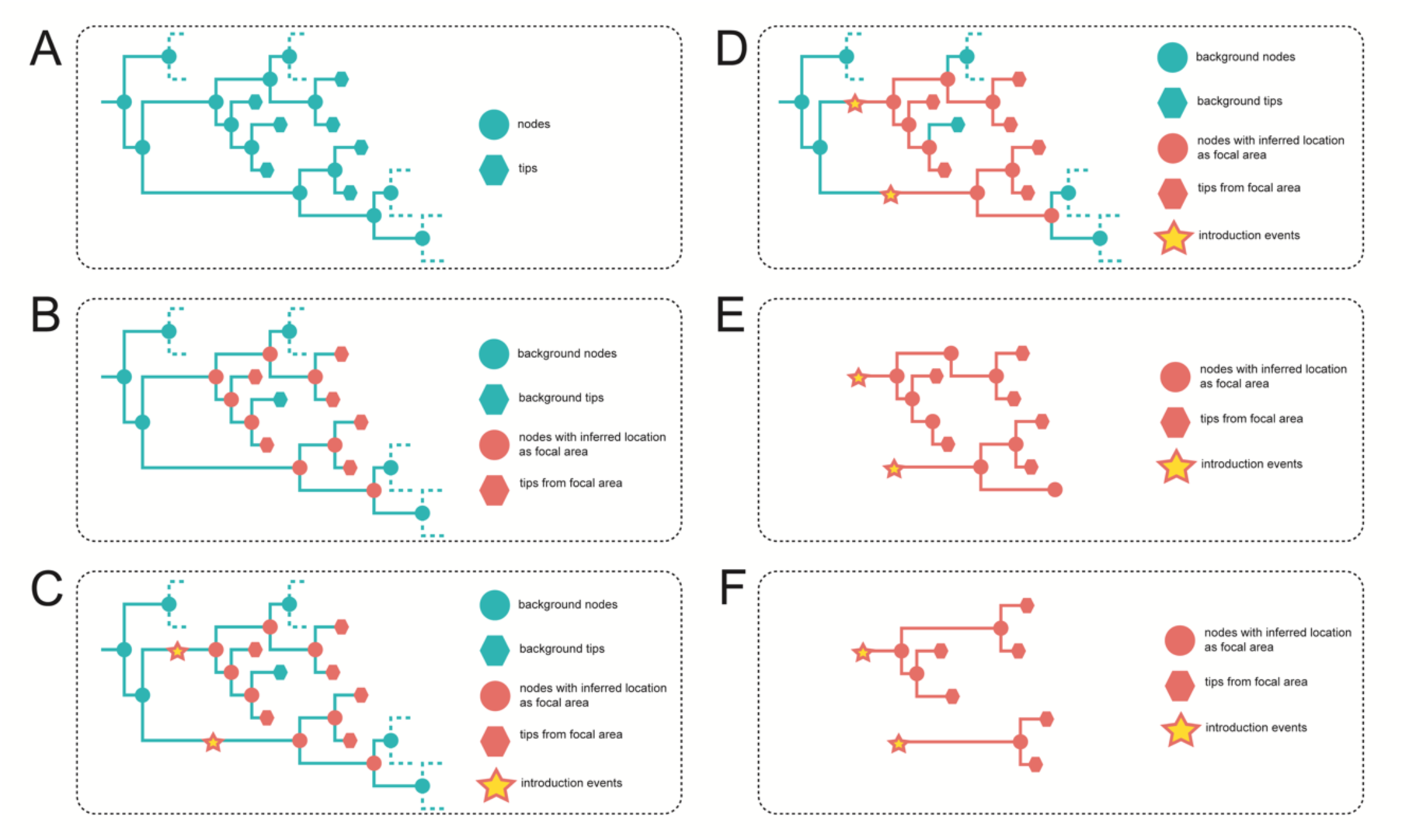
Outline of the Methodology for Extracting Locally Circulating Clusters. **A.** Phylogeny depicting isolates sampled from the focal area and background regions. **B.** Discrete trait analysis inferring geographic trait states for internal nodes. **C.** Identification of introduction events on branches connecting background nodes to nodes/tips from the focal area. **D.** Clade extension from introduction nodes to encompass all nodes and tips from the focal area. **E.** Overview of pseudo-clusters before refinement. **F.** Final locally circulating clusters obtained by collapsing branches connected to single-descent nodes.

We identified a total of 1,479 independent introduction events (95% highest posterior density [HPD]: 1,402–1,556). The sizes of the resulting circulating clusters were notably skewed (Figure 2A). Most introductions (909 events, 95% HPD: 853–968) resulted in singletons, while a few led to clusters exceeding 100 cases. Temporal analysis revealed that earlier introductions were more likely to produce larger clusters (Figure 2A). For instance, during EPI Week 17, 55.6% of introductions resulted in clusters larger than 10 cases, 33.3% in clusters smaller than 10, and 11.1% were singletons. By EPI Week 30, this distribution had shifted significantly, with 86.2% resulting in singletons, 13.8% forming clusters smaller than 10, and no clusters larger than 10.

**Figure 2.**
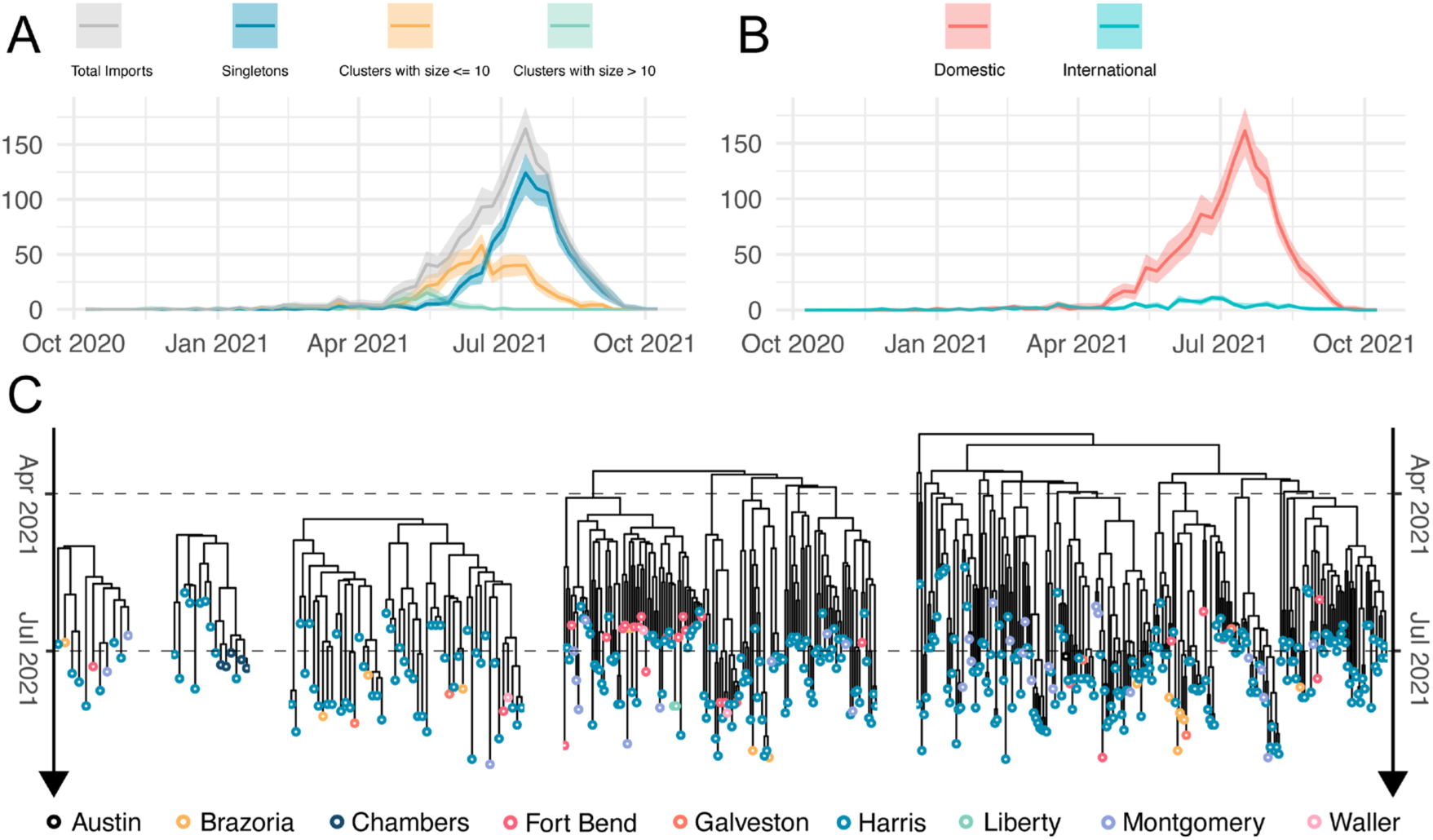
Dynamics of Viral Introduction into Greater Houston. **A.** Weekly frequency of viral introductions, with curves colored by the size of resulting local transmission lineages. Shaded regions represent the 95% HPD intervals. **B.** Weekly frequency of viral introductions, with curves colored to distinguish between domestic and international sources. Shaded regions represent the 95% HPD intervals. **C.** Local dispersal patterns in Houston illustrated using five randomly selected clusters. Tips are colored by the county where isolates were sampled.

We classified viral imports into Houston as either domestic or international based on their origin. At the onset of the outbreak, introductions were scattered from both sources. However, after late April, domestic imports rapidly increased and became the dominant type, while international imports peaked earlier (Figure 2B). A total of 1,359 domestic imports (95% HPD: 1,279–1,432) significantly outnumbered the 119 international imports (95% HPD: 109–132). To infer transmission parameters, we analyzed the size distribution of clusters, estimating the mean cluster size (r) and the dispersion parameter (k). Clusters resulting from international imports were associated with larger mean cluster size (r = 22.33) and greater variability in cluster sizes (k = 0.27) (Table 1).

**Table 1.**
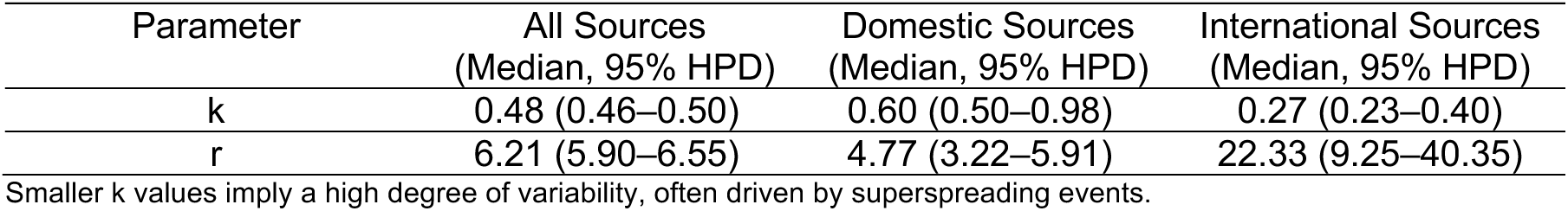
Characterizing Cluster Size Distributions with the dispersion parameter (k) and mean cluster size (r).

We selected 82 clusters, each containing more than 10 isolates, for further analysis of local dispersal. In total, these separate clusters included 6,455 sequences from the Greater Houston area, with the two largest clusters containing 2,159 and 1,878 sequences, respectively. Isolates from Harris County dominated the dataset and were present in all 82 clusters. To illustrate local dispersal patterns, we randomly selected five representative clusters, which are shown in Figure 2C.

### Phylogeny-Trait Correlation Among Locally Circulating Clusters

We investigated the correlations between phylogenetic structures and demographic traits, specifically age and sex, to better understand how these factors influence transmission dynamics (Figure 3A). Age groups were categorized as follows: infants and children (0–12 years), teenagers (13–18 years), young adults (19–35 years), middle-aged adults (36–55 years), and seniors (56 years and older). These age classifications were adapted from widely used demographic frameworks in public health and sociology.

**Figure 3.**
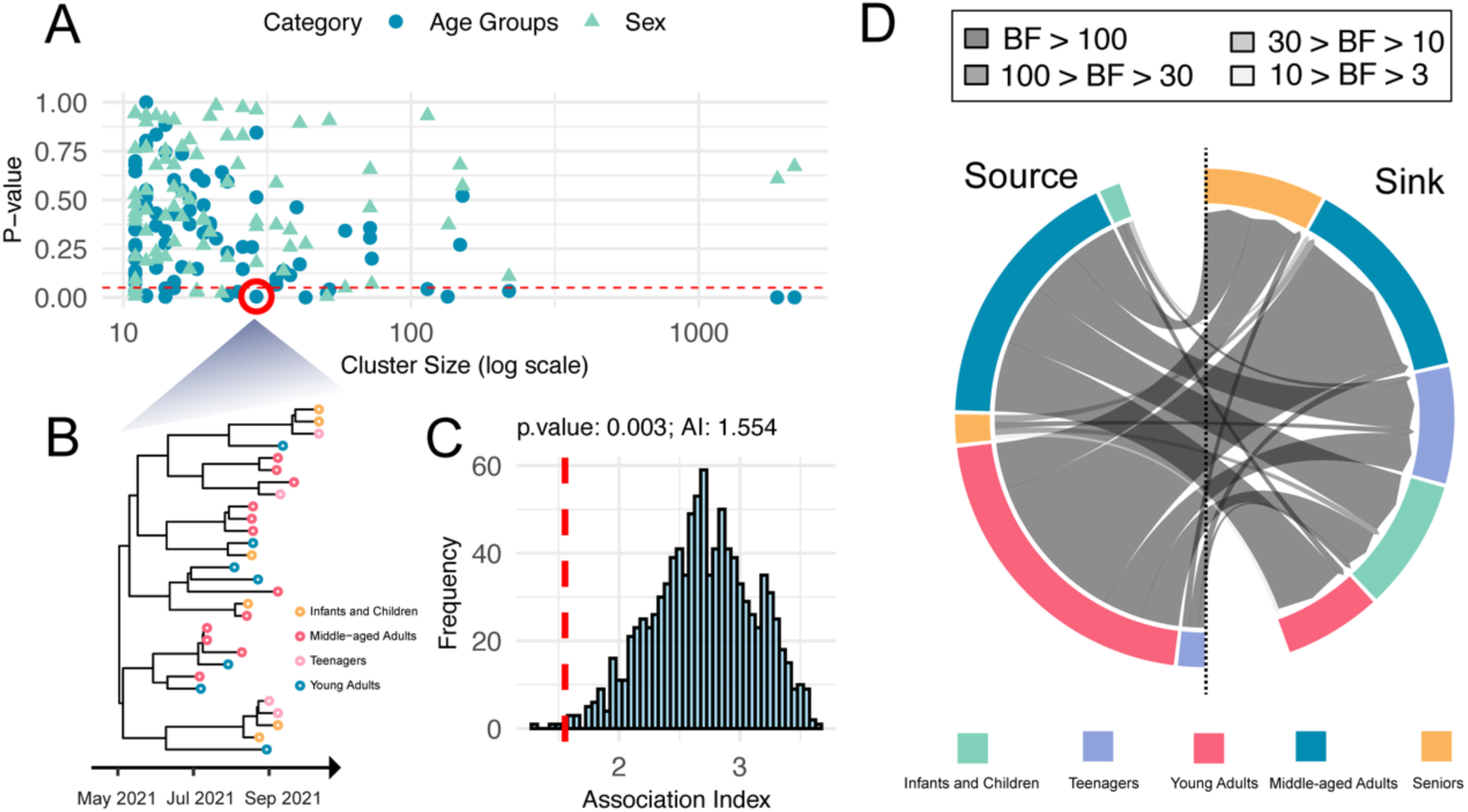
Demographic Determinants of Transmission. **A.** Association between 82 locally circulating clusters and demographic traits. Blue circles represent test results for age groups, and green triangles represent test results for sex groups. Shapes below the red line (p < 0.05) indicate clusters highly correlated with the corresponding traits. **B.** Detailed view of a highlighted cluster, with tips colored based on age groups. **C.** Null distribution of the association index for the highlighted cluster’s association test, with observed value indicated by a red dashed line. **D.** Discrete trait diffusion models for age groups, presented as circular charts. Chord thickness indicates the magnitude of transition rates, while color represents Bayes Factor (BF) support. Only transitions with BF > 3 from the discrete trait analysis are displayed.

For each locally circulating cluster (Figure 3B), we quantified phylogeny-trait correlations using the Association Index (27, 28). To evaluate the significance of these correlations, we generated null distributions of the Association Index by randomizing trait assignments across phylogeny tips 1,000 times (Figure 3C). This approach allowed us to perform the tip-trait association test. The null hypothesis assumed that traits were randomly associated with phylogenetic structures. A low p-value (p < 0.05) rejected this hypothesis, indicating a strong association and suggesting limited viral dispersal between different trait groups.

We tested 82 locally circulating clusters for associations with demographic traits (Figure 3A). Among these, 20 clusters exhibited significant correlations between age group traits and phylogeny (p < 0.05). In contrast, only 6 clusters showed significant correlations for sex traits. Overall, sex groups appeared more interspersed on the phylogeny, indicating that viral transmission is more constrained within age groups than within sex groups.

### Demographic Determinants of Transmission

Assuming (1) each local transmission cluster resulted from a single introduction into Houston, (2) all sequences within a cluster represented local transmission, and (3) each cluster was an independent observation of the same population process, we jointly estimated a single discrete trait model across all 82 circulating clusters to quantify viral dispersal among age groups. The analysis included 6,455 isolates distributed across the following age groups: 2,009 young adults, 1,995 middle-aged adults, 841 infants and children, 818 seniors, 771 teenagers, and 21 individuals of unknown age.

Our model (Figure 3D) identified young and middle-aged adults as the primary drivers of viral transmission. These groups were the sources of the highest transition rates, including: from young adults to middle-aged adults (4.813 transitions per year) and vice versa (2.216 transitions per year); from middle-aged adults to infants and children (1.895 transitions per year), seniors (1.518 transitions per year), and teenagers (1.516 transitions per year); and from young adults to seniors (1.459 transitions per year), teenagers (1.216 transitions per year), and infants and children (1.202 transitions per year). All transitions were strongly supported by Bayes Factors exceeding 100. A complete list of diffusion rates between age groups is provided in Table S1. Notably, the main sources of transmission to the vulnerable group—seniors—were teenagers, young adults, and middle-aged adults (BF > 100).

### Heterogeneous Dynamics of Viral Dispersal in Subregions of Greater Houston

We estimated a jointly fitted discrete model to reconstruct the geographic dispersal history of SARS-CoV-2 in Greater Houston. Categorizing location traits into 9 counties, the joint model (Figure 4A) identified 14 transitions decisively supported by Bayes Factors (>100). The highest transition rate was from Harris County to Fort Bend County (4.569 transitions per year), followed by transitions from Harris County to Montgomery County (3.292) and Harris County to Brazoria County (1.642). A detailed list of rates is provided in Table S2.

**Figure 4.**
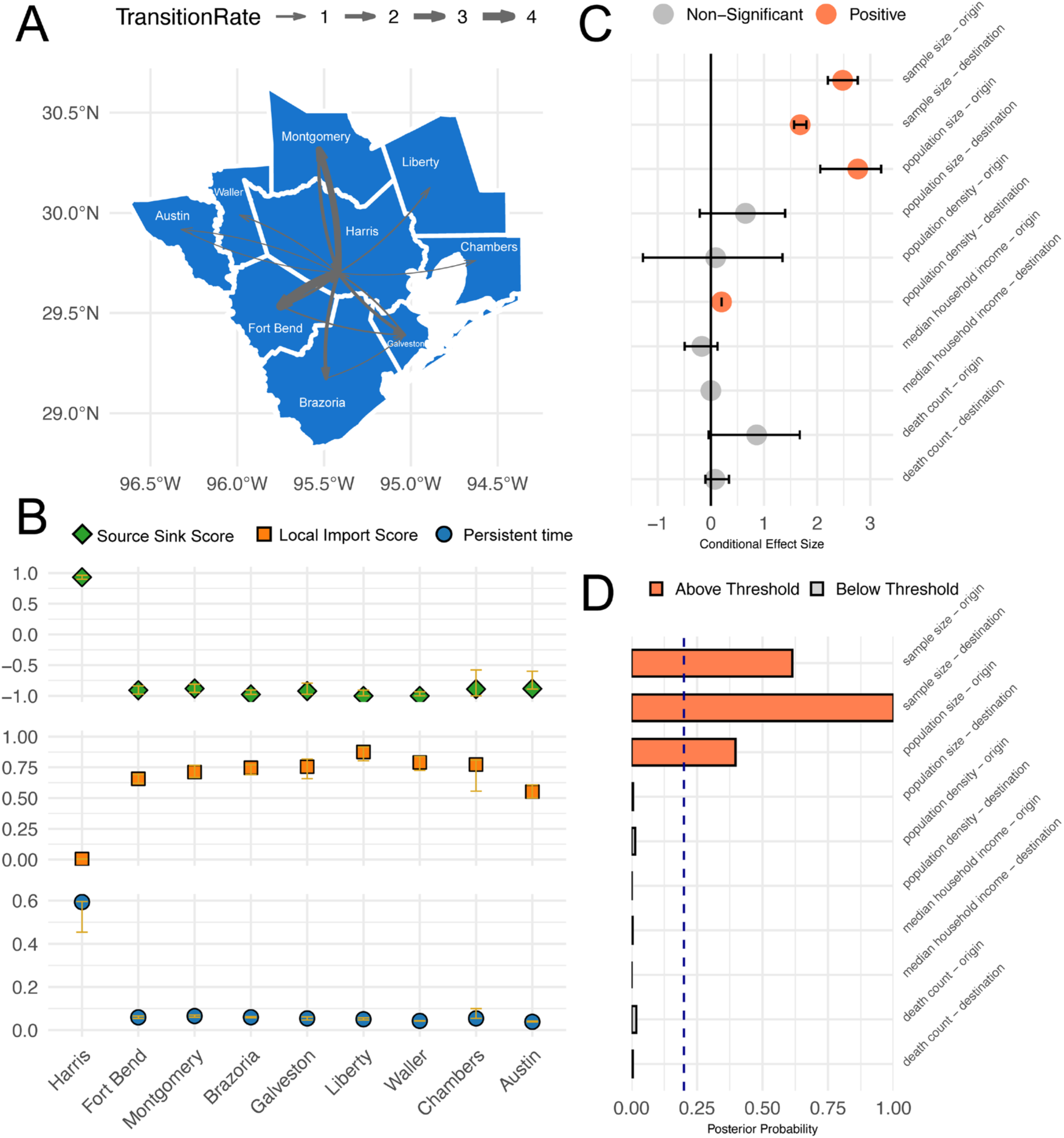
Distinct Transmission Patterns in Subregions of Greater Houston. **A.** Discrete phylogeographic reconstruction of the dispersal history across 9 counties. Arrow thickness indicates the magnitude of transition rates. All transitions shown on the map were decisively supported by Bayes factors (>100) **B.** Source Sink Scores, Local Import Scores, and Persistence Time across these subareas. In these bar charts, golden error bars represent the associated 95% HPD, providing a measure of uncertainty for each score. Source Sink Score ranges from 1 (viral source) to -1 (viral sink). Local Import Score ranges from 0 (epidemic is locally maintained) to 1 (epidemic relies on introduction). **C.** Conditional effect size of covariates within the generalized linear model (GLM). Conditional effect size represents the effect size of the variable coefficient given inclusion in the GLM. The black line represents a conditional effect size of 0, signifying little impact of the covariate on viral dispersal. **D.** Inclusion probabilities of covariates in the GLM, with a threshold set at a posterior probability of 0.2.

Discrete trait analysis yielded a sample of trees from the posterior distribution with resolved geographic states of internal nodes, enabling the calculation of metrics that summarize the transmission pattern of each subregion. These metrics include: (1) Source Sink Score (29), which identifies populations as either viral sources or sinks based on net viral flow weighted by outbreak size, (2) Local Import Score (29), which assesses the relative influence of viral introductions versus local transmission in driving the epidemic, and (3) Persistence Time (30–32), which is the average duration (in years) that a lineage remains within its sampled location, traced backward through the phylogeny. Harris County (Figure 4B) was identified as a key source of viral dispersal in Houston (Source Sink Score: 0.931, 95% HPD: 0.899–0.961). The epidemic in Harris County was predominantly driven by local transmission rather than viral introductions (Local Import Score: 0.004, 95% HPD: 0.002–0.006). Viral lineages also demonstrated longer persistence in Harris County (Persistence Time: 0.593 years, 95% HPD: 0.454–0.596 years). In contrast, viral sinks exhibited distinct transmission patterns, characterized by higher Local Import Score and shorter Persistence Time.

The discrete trait diffusion model was extended with a generalized linear model (GLM) to evaluate the association of demographic variables with transition rates between counties. We modeled SARS-CoV-2 transition rates as a function of 10 predictors: sample size, population size, population density, median household income, and accumulated death count for both origin and destination counties (Table S3). Three predictors were statistically supported (Figure 4C and Figure 4D): population size of the originating county (Conditional Effect Size: 2.763, 95% HPD: 2.058–3.202, Inclusion Probability: 0.387), sample size of the originating county (Conditional Effect Size: 2.479, 95% HPD: 2.201–2.763, Inclusion Probability: 0.614), and sample size of the destination county (Conditional Effect Size: 1.680, 95% HPD: 1.565–1.798, Inclusion Probability: 1).

## Discussion

The global spread of SARS-CoV-2 has triggered new outbreaks, but most cases arise from local transmission. Understanding local transmission dynamics quantitatively is essential for designing effective prevention strategies. In this study, we developed a novel algorithm to identify locally circulating clusters from phylogenetic data, incorporating contextual sequences. This approach allows us to assign each isolate from the focal area to a specific cluster, enabling a detailed analysis of the timing and frequency of introductions. The resulting clusters are essential for inferring local transmission patterns.

In collaboration with the Houston Health Department, we analyzed over 26,000 SARS-CoV-2 genomes and their associated metadata to study how the virus was introduced and spread within Greater Houston. This major metropolitan area is known for its demographic diversity. Our analysis covered the period between January and October 2021 and identified 1,479 independent introduction events (95% HPD: 1,402 to 1,556). Domestic origins were the predominant source of these introductions overall (Figure 2). We also examined how demographic structures influenced the spread of the virus. The tip-trait association test suggested that viral transmission was more restricted between age groups than between sexes (Figure 3A). Furthermore, discrete trait analysis modeled transmission between different age groups (Figure 3D). Lastly, we reconstructed the spatiotemporal spread of locally circulating clusters (Figure 4). Our analysis revealed that transmission dynamics varied across subregions of Greater Houston. For example, in Harris County, a key viral source, introductions made up a smaller proportion of new cases while local transmission chains were notably longer.

Our analysis quantitatively confirms that the Delta outbreak in Houston was driven by multiple independent introduction events. These introductions led to widespread local transmission, resulting in clusters of varying sizes. Earlier clusters were larger and more challenging to control. This pattern is consistent with previous findings on COVID-19 introductions in the UK (8, 33), New York City (34), and Denmark (5). At the onset of the outbreak, we observed that scattered introductions came from both domestic and international sources. However, after late April, domestic introductions surged and became dominant. Since we observed no clear predominance of international sources throughout the outbreak, we believe that Houston did not serve as a primary entry point for SARS-CoV-2 into the U.S. This contrasts with New York, California, and Florida, which have been identified as major entry points in previous research (4). Nevertheless, international introductions still had a significant impact, particularly during the early stages of the outbreak. These early international introductions often resulted in larger clusters and more sustained local transmission, placing a considerable burden on public health intervention.

Tip-trait association quantifies the degree to which viral phenotypic characters are correlated with shared ancestry, as represented by a viral phylogenetic tree (27). A common application of this phylogeny-trait correlation is to explore spatial structure (35); specifically, whether sequences group together in a phylogeny based on geographic location. In this study, we examine the correlations between phylogeny and population structures to better understand how demographic factors influence transmission dynamics. Human movements and interactions were generally more constrained by age group than by sex (36). For example, children are typically found in daycare centers or middle schools, teenagers in high schools, adults at their workplaces, and seniors in nursing homes. Our analysis on circulating clusters statistically supports that traits age groups are more tightly correlated with the tree topology, indicating more constrained transmission within these groups. Furthermore, we estimated the discrete trait model to describe the transmission between age groups. This analysis revealed that the primary sources of transmission to the vulnerable group—seniors—were teenagers, young adults, and middle-aged adults.

Previous reports showed the COVID-19 epidemic in Houston exhibited distinct patterns, including varying infection probabilities and hospitalization rates (22, 37). Here, we reconstructed spatial dispersal of SARS-CoV-2 across 9 subregions of Greater Houston using discrete trait analysis, applying the Source Sink Score, Local Import Score, and Persistent Time to characterize transmission patterns in each subregion. Through Bayesian phylogeographic inference, we were able to estimate the values of these metrics along with their highest posterior density intervals, providing a measure of confidence in our estimates. Our analysis revealed a consistent pattern across all subregions: regions with higher Source Sink Scores were associated with lower Local Import Scores and higher Persistent Times. This pattern aligns with previous analyses of viral transmission in Seattle (30), where a structured coalescent model (38) found that South King County exhibited longer persistence of local transmission compared to North King County, where external viral introductions drove a larger proportion of new cases. As the Source Sink Score provides evidence for whether a region functions as either a source or sink of viral transmission, these findings collectively suggest that a well-established and sustained local epidemic is crucial for a region to act as a source of pathogen spread to other areas. Targeted public health interventions in these identified source regions—such as temporary closures of schools, limitations on large public gatherings, enhanced testing and contact tracing, and increased access to healthcare resources—could not only mitigate local transmission but also have a broader impact by reducing the spread of the virus across the entire Greater Houston area. Such focused strategies can enhance the efficiency of outbreak control measures and allocate resources more effectively to areas with the greatest influence on regional transmission dynamics.

Our analysis is not without limitations. The algorithm for detecting clusters relies on incorporating background sequences from other locations. Differences in sampling ratios could bias this detection, raising questions about the sensitivity of this approach. For example, how does having fewer or more background samples affect cluster detection? Simulation studies could help address this by evaluating how the sampling ratios of background and focal sequences influence cluster detection accuracy. Additionally, simulations incorporating different reproductive numbers, generation times, and substitution rates could assess the algorithm’s performance under various pathogen dynamics.

## Materials and Methods

### SARS-CoV-2 Genomic Dataset

With the support of the Houston Health Department (HHD), we accessed a large dataset of SARS-CoV-2 genomes sampled in Houston, along with linked metadata, including ZIP code, age group, and sex. The first reported Delta variant case in Houston occurred in mid-April 2021. Our contextual dataset (non-Houston) was divided into two phases: Phase one included all worldwide sequences available in GISAID (www.gisaid.org) sampled before April 15, while Phase two sampled 1% of worldwide sequences available after April 15 (Figure S2). This sampling scheme balanced the need to include early sequences with the practical limits of handling a rapidly growing dataset.

The combined dataset of Delta sequences from Houston and contextual sources was then aligned to the reference genome (GenBank ID: NC_045512.2) using minimap2 v2.24 (39). We filtered out low-quality sequences with mapped completeness below 93% and trimmed alignments outside the reference coordinates 265:29674, padding with Ns to mask out the 3’ and 5’ UTRs. We then calculated the genetic base-pair differences between the alignments and the reference genome. For samples collected within the same Epi-Week, we excluded sequences with genetic differences greater than 3.0 standard deviations from the mean, aiming to preliminarily filter out those with a poor clock signal. In total, 26,138 alignments passed the filtering criteria. The contextual sequences were categorized as either domestic (excluding Houston) or international. Our dataset included 9,186 sequences from Houston, 5,334 from domestic sources, and 11,618 from international sources (Figure S3).

### Time-Scaled Phylogenetic Inference

Time-scaled phylogenies were inferred through the following steps: inferring an initial phylogenetic tree, calibrating it with time, and post-processing to refine the final tree. First, a maximum-likelihood phylogeny was estimated using IQ-TREE v2.3.2 (40), applying the HKY85 nucleotide substitution model with empirical base frequencies. The phylogeny was rooted using early samples from Wuhan (Wuhan-Hu-1/2019) to reflect the epidemic’s origins. Next, time calibration was performed using TreeTime v0.11.2 (41), which estimated an evolutionary rate of 0.0007 substitutions per site per year with r^2 = 0.35. A total of 102 tips with residuals greater than 3 interquartile distances from the residual distribution were flagged as outliers. Finally, the tree underwent post-processing steps. Outlier pruning was conducted using jclusterfunk v0.0.25 (42). The tree format was subsequently standardized using gotree v0.4.5 (43) to ensure compatibility for downstream analyses.

### Introduction Analysis

Before considering a discrete trait model to jointly reconstruct viral spread between locations and between age groups, we performed a preliminary discrete phylogeographic analysis (24) on the fixed time-scaled tree obtained in the previous step to identify independent introduction events. Tips of the tree were assigned locations of either ‘Houston’, ‘Domestic’, or ‘International’. After reconstructing the ancestral locations of the virus, we identified introduction events on branches connecting ‘non-Houston’ nodes (categorized as either ‘Domestic’ or ‘International’) to ‘Houston’ nodes or tips (Figure 1C). The introduction time was defined as the midpoint of these branches. Locally circulating clusters were then extracted by (1) extending pseudo-clusters from introduction events to encompass all nodes and tips assigned to ‘Houston’ and (2) refining these clusters by removing nodes with no descendants and collapsing branches connected to single-descent nodes (Figure 1D-F). This algorithm ensures that each isolate from Houston is assigned to a cluster. Phylogeny reading, editing, and writing were performed using the following R packages: ape 5.8 (44), treeio 1.20.2 (45), the tidytree 0.4.6 package (46), and ggtree 1.14.6 (47).

Based on the observed distribution of cluster sizes (assumed to follow a negative binomial distribution), we estimated the mean cluster size (r) and the dispersion parameter (k). Maximum likelihood estimation was performed using the ‘fitdistr’ function from the MASS package in R (48).

We simulated 5 Markov chains of 20 million states each, with a burn-in of 4 million states, using BEAST v1.10.5 (49) with XML input files generated using custom R scripts. Trees were sampled every 160,000 states, resulting in an empirical tree set size of 500. The convergence and mixing of all relevant parameters were inspected using Tracer 1.7 (50). We analyzed the posterior tree sets to infer 95% HPD intervals for the weekly count of introduction events, the mean cluster size (r) and the dispersion parameter (k). A representative tree was selected from the posterior tree set, matching the posterior median for total introductions (Figure S4). From this tree, we extracted 82 locally circulating clusters, each comprising more than 10 isolates.

### Tip-Trait Association Test

For each locally circulating cluster, we used the Association Index to quantify phylogeny-trait correlations. To assess the significance of these correlations, we generated null distributions of the Association Index by randomizing trait assignments on the tips 1,000 times, enabling us to perform the tip-trait association test. All scripts for this test were bundled into an R package named TTAT, which is publicly available on GitHub at https://github.com/leke-lyu/TTAT.

### Jointly Fitted Discrete Trait Model

We modeled the evolution of discrete traits on local clusters using a continuous-time Markov chain (CTMC), analogous to standard approaches for modeling molecular sequence evolution (24). Assuming that all 82 circulating clusters shared the same population process, we jointly estimated two separate transition rate matrices (34, 51, 52): one describing transitions between age groups and another describing transitions between counties. The age group model included 5 distinct categories: infants and children, teenagers, young adults, middle-aged adults, and seniors. We assumed a non-reversible transition model (53) consisting of 20 separate rate parameters, each augmented with a binary indicator variable to perform Bayesian stochastic search variable selection (BSSVS) (54) . Similarly, the geographic model analyzed transitions between 9 counties, applying a non-reversible transition model with 72 separate rate parameters, each accompanied by a BSSVS indicator variable.

For both models, a Poisson prior with a mean of 1 and an offset of 0 was used for the total number of non-zero rates. XML configuration files for the BEAST runs were generated using custom R scripts. We performed five independent chains, each consisting of 100 million states with a burn-in period of 20 million states. Trees were sampled every 800,000 states, yielding an empirical tree set of 500 samples per cluster. Convergence and mixing were verified using Tracer.

The geographic model was further extended with a generalized linear model (GLM) (25), where viral diffusion rates among counties served as the outcome in a log-linear combination of epidemiological predictors, regression coefficients, and indicator variables for BSSVS. Population size, population density, and median household income for all nine counties were obtained from the U.S. Census Bureau. Accumulated SARS-CoV-2 death counts were obtained from the Texas COVID-19 Surveillance Archives (55).

### Posterior Processing

The jointly fitted discrete trait model reconstructed the geographic states of internal nodes for 82 locally circulating clusters. Given these posterior tree sets, we estimated the following epidemiological metrics that summarize the transmission pattern of each subregion:

A. Source Sink Score: This metric, ranging between -1 and 1, measures net viral exports, weighted by outbreak size. A score approaching 1 suggests that the region primarily functions as a viral source. Conversely, a score nearing -1 indicates that the region predominantly acts as a viral sink.
B. Local Import Score: Ranging from 0 to 1, this metric measures the fraction of introductions relative to the total count of new cases in a region. A score near 1 indicates that introductions predominate, while a score near 0 suggests that local transmissions dominate, indicating that the epidemic is primarily sustained locally.
C. Persistence Time: This metric measures how long a viral lineage circulates in a region. It is calculated by tracing the number of days it takes for a tip to move from its sampled location, going backward up the phylogeny until the node location differs from the tip location.

## Data Availability

All data produced in the present study are available upon reasonable request to the authors.

## Acknowledgments

This work has been funded in part from the National Institute of Allergy and Infectious Diseases, a component of the NIH, Department of Health and Human Services, under contract no. 75N93021C00018 (NIAID Centers of Excellence for Influenza Research and Response, CEIRR) and Centers for Disease Control and Prevention, Department of Health and Human Services, under contracts 75D30121C10133 and NU50CK000626. Scripts used to generate the results in the Texas case study are publicly available at https://github.com/leke-lyu/jointEstimation. We acknowledge the GISAID contributors (acknowledgment table of genomes used is provided on our GitHub repository) for sharing genomic data.

