## Supporting Information for "Tracing SARS-CoV-2 Clusters Across Local Scales Using Genomic Data"

**This PDF file includes:**

Figures S1 to S4

Tables S1 to S3

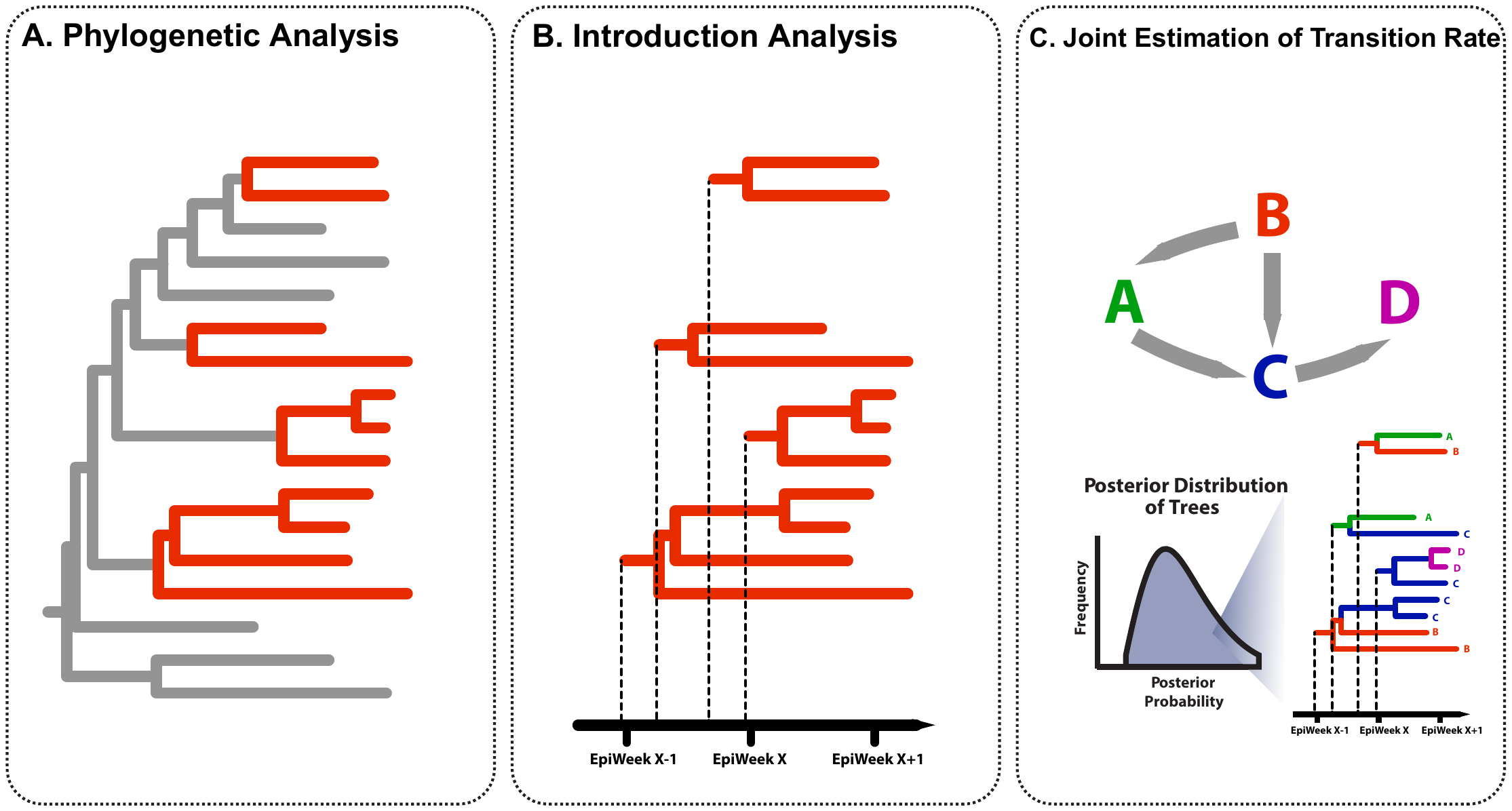

Fig. S1. Conceptual Workflow of Large-Scale Genomic Epidemiology Analysis. Phylogenetic Analysis infers the phylogeny representing the evolutionary history of the isolated sampled from the focal region within a global context. Introduction analysis estimates the timing of viral introductions and identifies locally circulating clusters. The Joint Fit Model infers transition rates under a unified transition matrix.

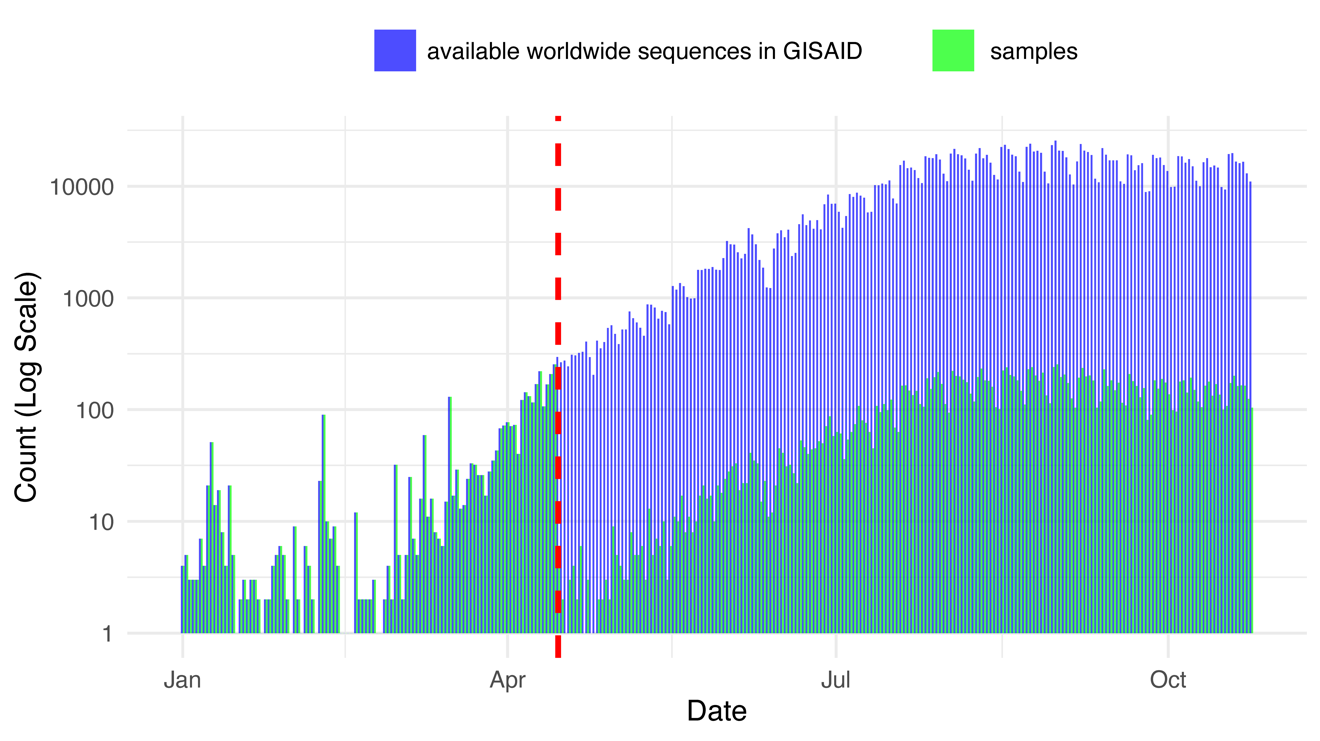

Fig. S2. Sampling scheme for contextual sequences. The contextual sequence dataset includes all worldwide sequences available in GISAID ([www.gisaid.org](http://www.gisaid.org/)) sampled before April 15, as well as 1% of the worldwide sequences available after April 15.

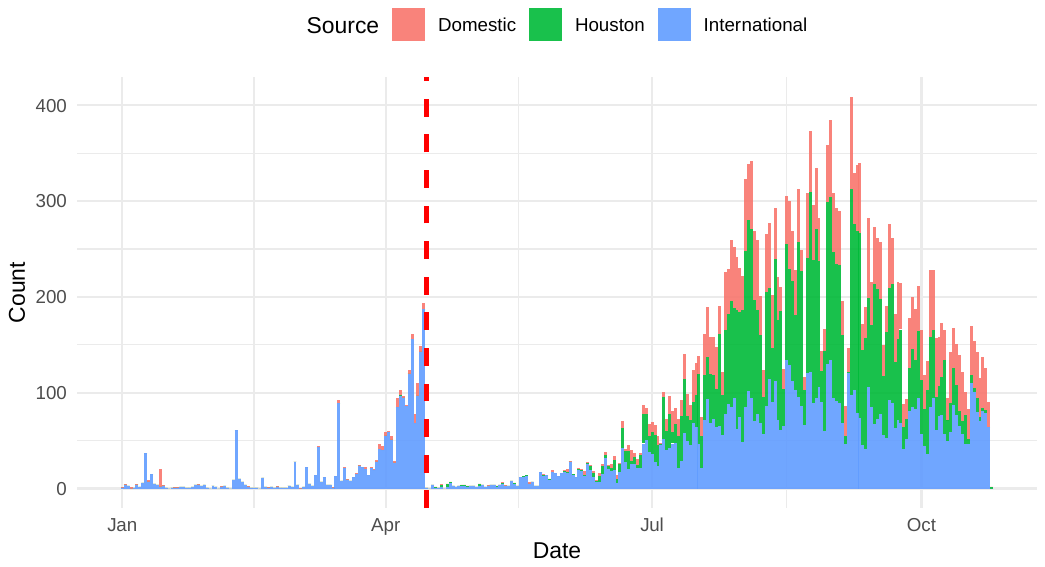

Fig. S3. Sampling date distribution of the complete genome dataset. The bars in the plot represent the daily count of isolates, categorized by source (international, domestic, and Houston) and distinguished by color.

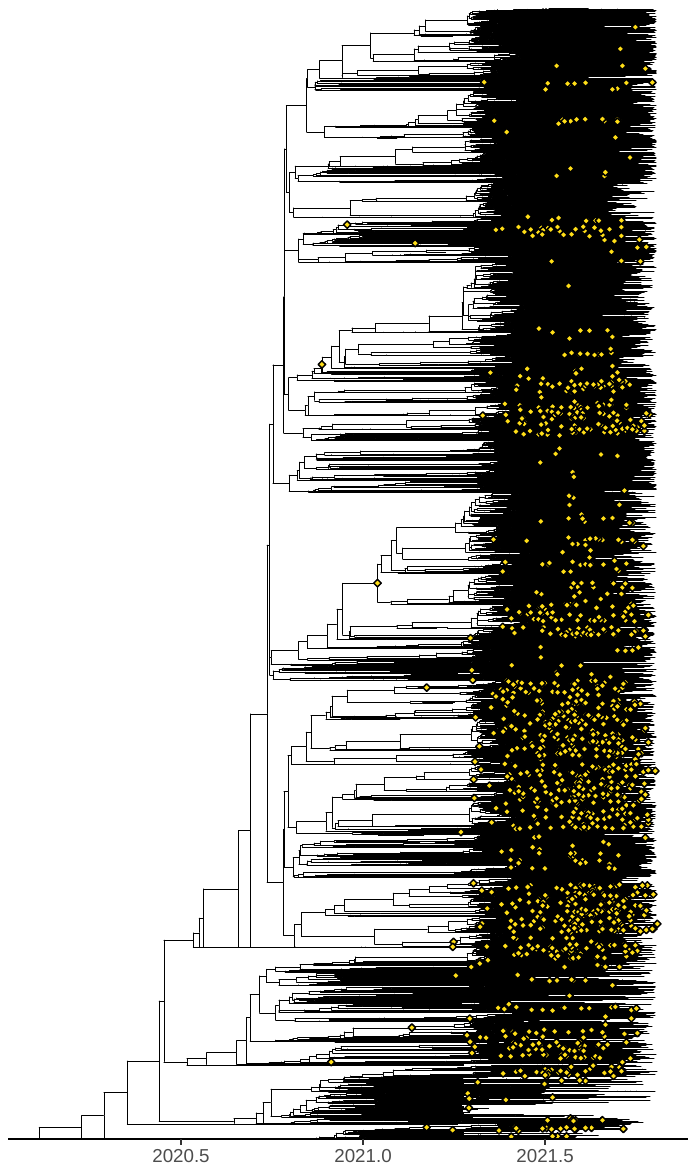

Fig. S4. The complete phylogeny of >26,000 isolates. A representative tree was selected from the posterior tree set, matching the posterior median for total importations. Identified introduction events to Houston are highlighted as yellow diamonds on the branches.

Table S1. Transmission rates between age groups. Age groups were categorized as follows: infants and children (0–12 years), teenagers (13–18 years), young adults (19–35 years), middle-aged adults (36–55 years), and seniors (56 years and older). All transmissions shown here have an inclusion probability > 0.5. The Bayes factors are provided for each rate.

| From | To | Transition Rate | BF |
| --- | --- | --- | --- |
| Young Adults | Middle-aged Adults | 4.813 | > 10,000 |
| Middle-aged Adults | Young Adults | 2.216 | > 10,000 |
| Middle-aged Adults | Infants and Children | 1.895 | > 10,000 |
| Middle-aged Adults | Seniors | 1.518 | > 10,000 |
| Middle-aged Adults | Teenagers | 1.516 | > 10,000 |
| Young Adults | Seniors | 1.459 | > 10,000 |
| Young Adults | Teenagers | 1.216 | > 10,000 |
| Young Adults | Infants and Children | 1.202 | > 10,000 |
| Teenagers | Middle-aged Adults | 0.314 | 3258.597 |
| Seniors | Middle-aged Adults | 0.234 | 51.837 |
| Teenagers | Seniors | 0.229 | > 10,000 |
| Infants and Children | Young Adults | 0.214 | > 10,000 |
| Seniors | Infants and Children | 0.208 | 72.596 |
| Seniors | Teenagers | 0.193 | 1915.479 |
| Infants and Children | Teenagers | 0.180 | 1479.402 |
| Teenagers | Infants and Children | 0.163 | 122.195 |
| Infants and Children | Middle-aged Adults | 0.156 | 13.449 |
| Seniors | Young Adults | 0.138 | 5.430 |

Table S2. Transmission rates between counties. All transmissions shown here have an inclusion probability > 0.5. The Bayes factors are provided for each rate.

| From | To | Transition Rate | BF |
| --- | --- | --- | --- |
| Harris | Fort Bend | 4.569 | > 10,000 |
| Harris | Montgomery | 3.292 | > 10,000 |
| Harris | Brazoria | 1.642 | > 10,000 |
| Montgomery | Harris | 1.455 | > 10,000 |
| Harris | Galveston | 1.373 | > 10,000 |
| Fort Bend | Harris | 1.147 | > 10,000 |
| Galveston | Harris | 0.546 | > 10,000 |
| Fort Bend | Galveston | 0.460 | 166.107 |
| Harris | Liberty | 0.423 | > 10,000 |
| Harris | Waller | 0.301 | > 10,000 |
| Brazoria | Galveston | 0.214 | > 10,000 |
| Harris | Chambers | 0.170 | > 10,000 |
| Harris | Austin | 0.169 | > 10,000 |
| Chambers | Harris | 0.166 | 9.614 |
| Austin | Harris | 0.139 | 7275.103 |

Table S3. Predictors used in the GLM

| County | Case | Population | Population Density | Median Household Income | Death |
| --- | --- | --- | --- | --- | --- |
| Austin | 2,140 | 30,167 | 46.7 | 68,630 | 43 |
| Brazoria | 36,372 | 372,031 | 274.0 | 86,197 | 625 |
| Chambers | 4,606 | 46,571 | 78.0 | 102,146 | 48 |
| Fort Bend | 72,153 | 822,779 | 955.1 | 105,583 | 603 |
| Galveston | 40,631 | 350,682 | 926.8 | 75,947 | 505 |
| Harris | 398,729 | 4,731,145 | 2777.3 | 68,748 | 6152 |
| Liberty | 7,280 | 91,628 | 79.1 | 61,880 | 265 |
| Montgomery | 50,725 | 620,443 | 595.6 | 95,241 | 855 |
| Waller | 3,886 | 56,794 | 110.6 | 75,223 | 66 |
